# Educational Mobility, the Pace of Biological Aging, and Lifespan in the Framingham Heart Study

**DOI:** 10.1101/2023.11.04.23298091

**Authors:** G.H. Graf, A.E. Aiello, A. Caspi, M. Kothari, H. Liu, T.E. Moffitt, P. Muennig, C.P. Ryan, K. Sugden, D.W. Belsky

## Abstract

**Importance:** People who complete more education live longer lives with better health. New evidence suggests that these benefits operate through a slowed pace of biological aging. If so, measurements of the pace biological aging could offer intermediate endpoints for studies of how interventions to promote education will impact healthy longevity.

**Objective:** To test the hypothesis that upward educational mobility contributes to a slower pace of biological aging and increased longevity.

**Design:** Prospective cohort study.

**Setting:** We analyzed data from three generations of participants in the Framingham Heart Study: the Original cohort, enrolled beginning in 1948, the Offspring cohort, enrolled beginning in 1971, and the Gen3 cohort, enrolled beginning in 2002. Follow-up is on-going. Data analysis was conducted during 2022-2023 using data obtained from dbGaP (phs000007.v33).

**Participants:** We constructed a three-generation database to quantify intergenerational educational mobility. We linked mobility data with blood DNA methylation data collected from the Offspring cohort in (2005-2008) (n=1,652) and the Gen3 cohort in 2009-2011 (n=1,449). These n=3,101 participants formed our analysis sample.

**Exposure:** We measured educational mobility by comparing participants’ educational outcomes with those of their parents.

**Outcomes:** We measured the pace of biological aging from whole-blood DNA-methylation data using the DunedinPACE epigenetic clock. For comparison purposes, we repeated analysis using four other epigenetic clocks. Survival follow-up was conducted through 2019.

**Results:** Participants who were upwardly mobile in educational terms tended to have slower DunedinPACE in later life (r=-0.18, 95% CI [-0.23,-0.13], p<0.001). This pattern of association was similar across generations and held in within-family sibling comparisons. 402 Offspring-cohort participants died over the follow-up period. Upward educational mobility was associated with lower mortality risk (HR=0.89, 95% CI [0.81,0.98] p=0.014). Slower DunedinPACE accounted for roughly half of this association.

**Conclusions and Relevance:** Our findings support the hypothesis that interventions to promote educational attainment will slow the pace of biological aging and promote longevity. Epigenetic clocks, like DunedinPACE, have potential as near-term outcome measures of intervention effects on healthy aging. Experimental evidence is needed to confirm findings.

## INTRODUCTION

People who complete more years of schooling tend to live longer, healthier lives. This educational gradient is thought to arise through improvements in socioeconomic resources and resulting access to health services, health-promoting social networks and communities, and healthy behaviors (1,2). Educational gradients are apparent in nearly every organ system and aging-related disease, including heart disease, diabetes, cancer, Alzheimer’s disease, etc. (3–11); people with higher levels of education experience a lower prevalence of aging-related disease and later age of onset of disease. This evidence of more rapid decline across organ systems suggests an overall acceleration of the pace of biological aging.

Biological aging refers to a set of processes characterized by an accumulation of molecular changes or "hallmarks" that progressively undermine the integrity and resilience capacity of our cells, tissues, and organs as we grow older (12,13). We recently developed a novel method to quantify the pace of biological aging in humans. Our approach used longitudinal phenotyping of multiorgan system decline to derive a DNA-methylation blood test measurement of the pace of biological aging, DunedinPACE: **P**ace of **A**ging **C**alculated in the **E**pigenome (14).

We recently showed that DunedinPACE is accelerated in individuals with low levels of education, and slowed in those with higher levels of education (15,16). In this study, we build on these observations to test the hypothesis that higher educational attainment promotes longevity by slowing the pace of aging. Because educational attainment is influenced by both social factors and genetic inheritance (17–20), we focused our analysis on educational mobility, i.e., differences in educational achievements of children relative to their parents. We further conducted analysis of sibling differences to address potential confounding by other factors shared within a family (21).

We measured participants’ educational mobility by linking educational records across the original cohort of Framingham Heart Study ("Original” cohort), the cohort formed from the Original cohort’s children and their spouses (“Offspring” cohort), and the cohort formed from the Offspring cohort’s children and their spouses (“Gen3” cohort). This procedure allowed us to compute educational mobility for members of the Offspring and Gen3 cohorts. For these participants, we measured pace of aging from blood DNA-methylation data using the DunedinPACE algorithm. For Offspring cohort participants, we measured longevity outcomes over 15 years of follow-up from the time of DNA methylation measurement. Our analysis proceeded in two steps. We first tested associations between educational mobility and biological aging among participants for whom data on educational mobility and blood DNA methylation were available. We then tested the extent to which mobility-mortality associations were mediated through the pace of biological aging.

## METHODS

### Data and Participants

The Framingham Heart Study (FHS) is an ongoing observational cohort study first initiated in 1948. Since study initiation, two additional cohorts have been recruited, consisting of the children and grandchildren of the original Framingham cohort and their spouses. DNA methylation data are available from blood tests administered during the Offspring cohort’s 8^th^ Exam (2005-2008) and the Gen3 cohort’s second exam (2009-2011).

We analyzed data from n=14,106 participants across all generations for whom education data were available. Our DNA-methylation analysis sample consisted of participants who (1) provided data on their own educational attainment; (2) who could be linked to educational data from at least one parent; and (3) had provided a blood sample for DNA methylation analysis. This sample included 1,652 members of the Offspring cohort (46% male, mean age at DNA-methylation measurement=66, SD=9) from 1,025 families and 1,449 members of the Gen3 cohort (48% male, mean age at DNA-methylation measurement=45, SD=8) from 552 families. Participant flow diagrams are shown in **Fig. 1**.

**Figure 1.**
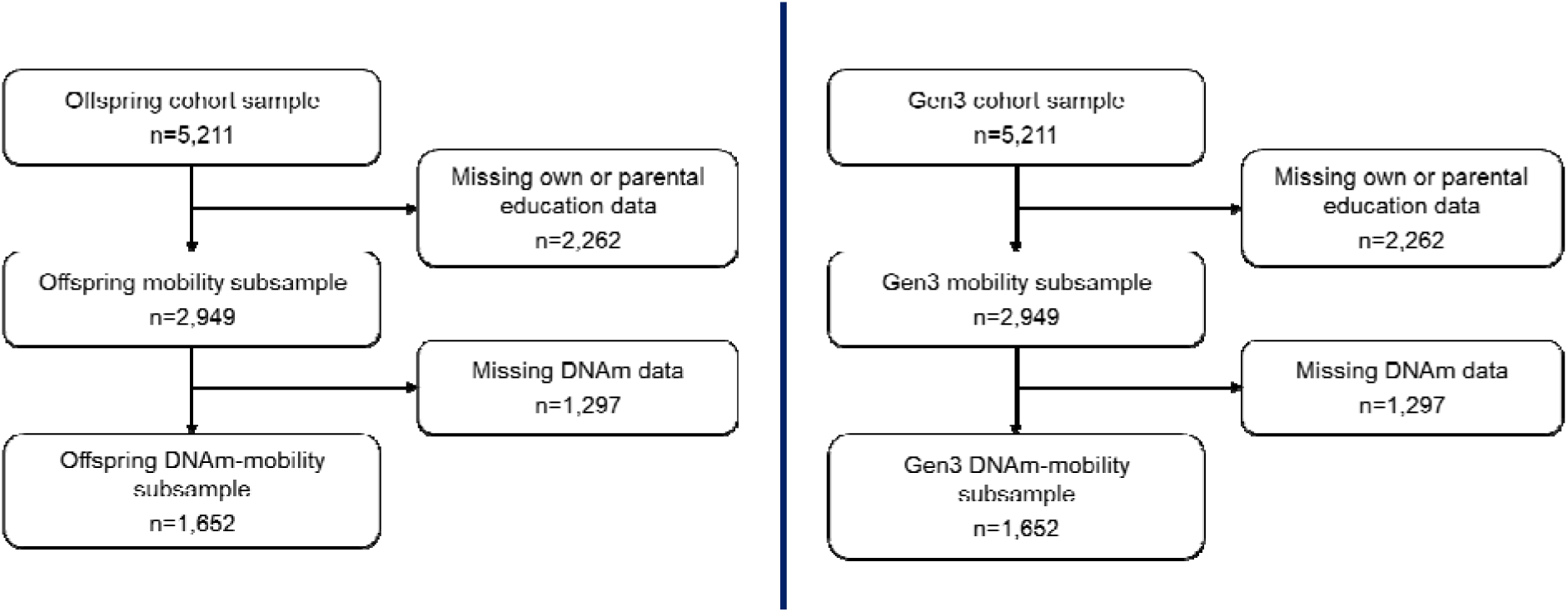
Offspring and Gen3 participant flow diagrams. The figure shows how the final analytic samples were developed from the larger set of all participants in the Offspring (left) and Gen3 (right) Framingham cohorts (combined n=3,101).

### Educational mobility

#### Educational attainment

We harmonized measures of educational attainment across FHS cohorts by converting reported levels of educational attainment completed years of education, following the method used by Liu and colleagues (18).

Within each of the three FHS cohorts, participants span a wide age range. For example, for participants in our Offspring cohort sample, ages at DNA-methylation (DNAm) measurement ranged from 40-92, corresponding to birth years spanning the early through mid-20th Century. More recent birth cohorts have generally experienced social and policy environments that encouraged higher educational attainment compared to their earlier-born counterparts (22). To adjust for this temporal trend in educational attainment, we converted each participant’s completed years of education into z-scores standardized to gender and 5-year birth cohort. A comprehensive account of educational attainment by birth cohort, including the methodology employed for data harmonization and standardization, can be found in the **Supplemental Methods**.

#### Educational mobility

We linked records of participants and their parents. Participants whose parents had higher levels of educational attainment tended to have higher educational attainment themselves (Pearson’s r=0.35; **Supplemental Fig. S1**). We computed mobility values using residualized-change scores, which quantify mobility as the difference between a participant’s educational attainment and the attainment expected based on the educational levels of their parents, and difference scores, which quantify mobility as the raw difference between parental and offspring educational attainment. Details of both approaches have been discussed in our previous work (16). Both metrics are denominated in sex- and birth-cohort-standardized units of education. On average, for both the residualized-change and difference-score methods, each unit of mobility corresponded to approximately two years of education.

### Biological Aging

Whole-genome DNA methylation (DNAm) profiles were obtained from dbGaP (phs000724.v10.p14). Details of the data are reported in the **Supplemental Methods.**

#### DunedinPACE

Biological aging is the progressive loss of integrity and resilience capacity in our cells, tissues, and organs that occurs with advancing chronological age (23,24). Pace of Aging is a phenotype reflecting the rate at which these biological changes occur (25). We quantified pace of biological aging using the DunedinPACE DNA methylation algorithm (14). The measure was constructed from analysis of 20-year longitudinal change in 19 biomarkers of organ-system integrity in the Dunedin Study 1972-3 birth cohort at ages 26, 32, 38, and 45 (26). The longitudinal Pace of Aging measure was then distilled into a single-timepoint DNA methylation blood test using elastic-net regression (27). In diverse cohorts in the US and around the globe, DunedinPACE is associated with incident morbidity and disability, survival, and a range of socioenvironmental exposures including educational attainment (14,15,28–36). DunedinPACE values were computed following the procedure described by Belsky and colleagues (14) using code available on GitHub (https://github.com/danbelsky/DunedinPACE).

#### Other epigenetic clocks

Other candidate measures of aging can be computed from DNA-methylation data. For comparison purposes, we repeated our analysis using four alternative “epigenetic clocks” which are widely studied in the literature and which we have reported on in prior studies: the Horvath, Hannum, PhenoAge, and GrimAge clocks (37–40). These clocks were calculated using the online calculator hosted by the Horvath Lab (https://dnamage.genetics.ucla.edu/new). Biological-age advancement was calculated by residualizing biological age values for chronological age at the time of DNA measurement.

### Survival

Details of FHS survival and mortality follow-up are reported elsewhere (41). Briefly, FHS conducts continuous mortality follow-up for all study participants. Date and cause of death are recorded for each participant based on hospital records, death certificates, and next-of-kin interviews. The present study included mortality data accumulated through 2019 (mean follow-up from DNA methylation baseline=12 years).

### Analysis

We tested associations of educational mobility with DunedinPACE using linear regression models. We used generalized estimating equations to account for non-independence of observations of individuals within nuclear families (42). We conducted within-family analysis comparing sibling differences in educational attainment with sibling differences in DunedinPACE using fixed effects regression (43). We tested associations of educational mobility and DunedinPACE with survival time using Cox proportional hazard regression models. Mediation analysis was conducted using the CMAverse package (44) in the R programming environment (45) following the approach described by Valeri and Vanderweele (46). All models were adjusted for age and sex.

## RESULTS

Participant characteristics are reported in **Table 1**. FHS participants with data on education and educational mobility were similar to the overall DNA-methylation sample.

**Table 1.** Characteristics of DNA-methylation, education, and mobility samples. The table provides information on the composition of our analytic sample (n=3,101, Offspring n=1,652, Gen3 n=1,449). The sample includes all individuals all individuals who provided DNA-methylation data, who reported their own educational attainment levels, and for whom educational attainment data was available for at least one parent.

|  | Analytic Sample (n=3936) | Offspring sample (n=3874) | Gen3 Sample (n=3101) |
| --- | --- | --- | --- |
| No. families | 1577 | 1025 | 552 |
| No. individuals | 3101 | 1652 | 1449 |
| Age (Years) | 56.14 (13.25) | 65.57 (9.22) | 45.38 (7.83) |
| Male | 47% | 46% | 48% |
| Died | 14% | 24% |  |
| Education (Years) | 14.97 (2.13) | 14.74 (2.31) | 15.24 (1.88) |
| Parental Education (Years) | 13.58 (2.7) | 12.35 (2.45) | 14.98 (2.26) |
| Ed. Mobility (Delta, Years) | 1.24 (2.47) | 1.99 (2.48) | 0.4 (2.16) |
| Ed. Mobility (Resid) | 0.04 (0.88) | 0.09 (0.88) | -0.02 (0.89) |
| DunedinPACE | 1.06 (0.12) | 1.08 (0.12) | 1.03 (0.11) |

### FHS participants who were upwardly mobile in their educational attainment experienced slower pace of biological aging

Participants who were upwardly mobile had slower DunedinPACE than those who were downwardly mobile (for residualized-change mobility, Offspring cohort Cohen’s d=-0.18, 95%CI=[-0.23,-0.13], p<0.001; Gen3 cohort Cohen’s d=-0.22, 95%CI=[-0.27,-0.16], p<0.001; for difference-score mobility, Offspring cohort Cohen’s d=-0.07, 95%CI=[-0.11,-0.03], p<0.0015; Gen3 cohort Cohen’s d=-0.08, 95%CI=[-0.13,-0.03], p<0.0012; **Supplemental Table S1, Fig. 2**). As a sensitivity analyses, we tested consistency of associations across participants who were born into lower- and higher-educated families to evaluate whether returns to educational mobility were concentrated at one end of the socioeconomic continuum. Effect-sizes were similar across strata of parental education (**Supplemental Fig. S2**). In addition, effect-sizes for educational mobility were comparable in the Offspring and Gen3 cohorts, suggesting consistent returns to relative educational mobility over time (**Fig. 2**). In comparative analysis of other epigenetic clocks, associations were weaker and not statistically different from zero for the Horvath, Hannum, and PhenoAge clocks. Results for the GrimAge clock, which was developed within the Framingham Heart Study, were similar to those for DunedinPACE. Full results are reported in **Supplemental Table S1**.

**Figure 2.**
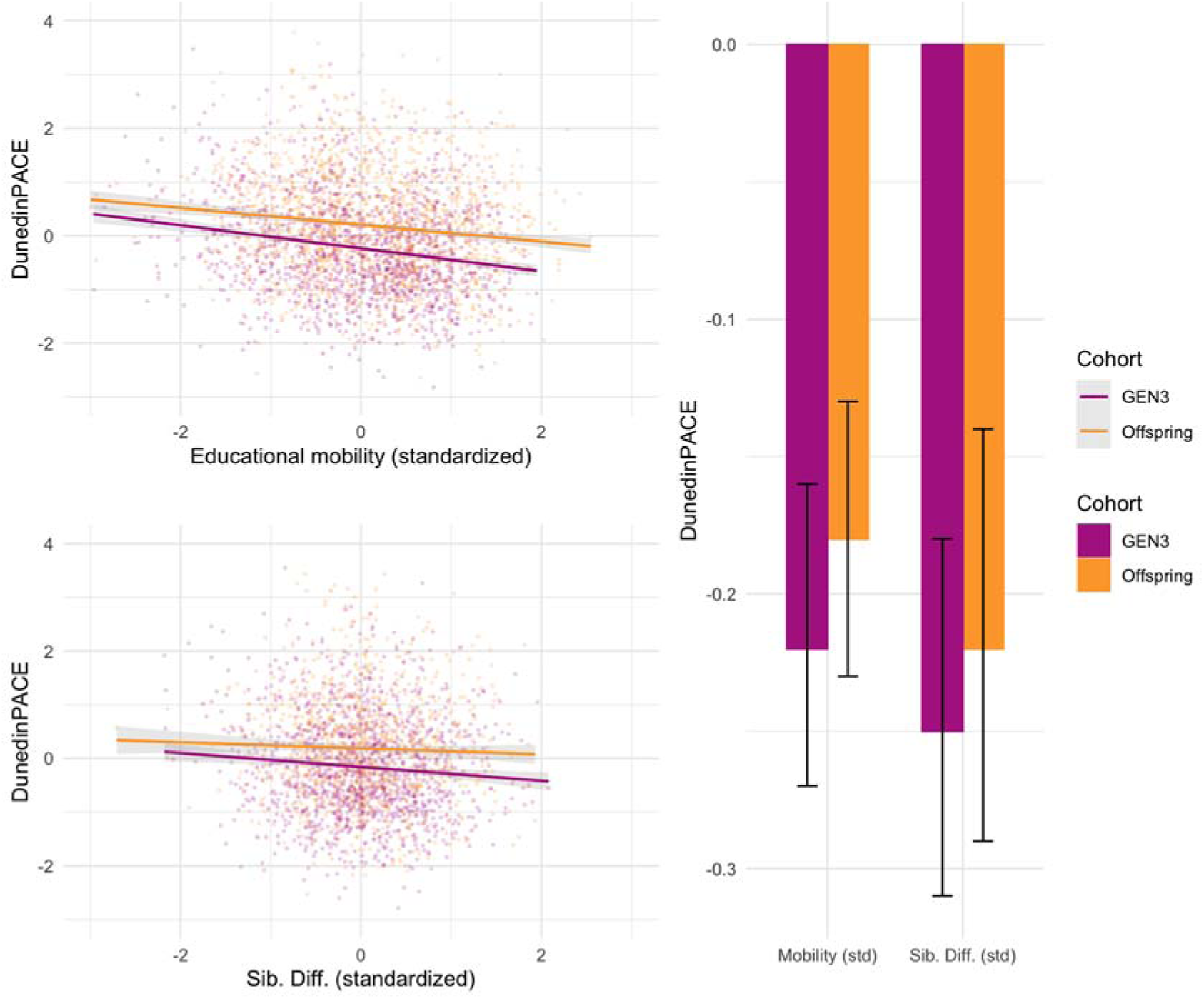
Association of educational mobility with DunedinPACE. The figure shows associations of educational mobility with faster pace of aging. The left side of the figure shows scatterplots and fitted regression slopes for the Offspring (orange) and Gen3 (purple) cohorts. The right side of the figure shows effect-estimates and 95% confidence intervals from regression analysis with covariate adjustments for participant age and sex. Effect-sizes are reported in standard-deviation units of DunedinPACE per standard deviation unit of educational mobility (interpretable on a Pearson r scale). Results are shown for analysis of educational mobility computed as the difference in education between children and their parents (top scatterplot, left-side set of bars) and for sibling differences in educational outcomes (bottom scatterplot, right-side set of bars).

### Among siblings, higher educational attainment was associated with slower pace of aging

To the extent that there are social or environmental factors that affect both educational mobility and aging trajectories, our results may overstate the impacts of mobility on healthy aging. To address this potential confound, we repeated our analysis within families. For participants with a sibling in the data (n=2,437; Offspring Cohort n=1,096; Gen3 Cohort n=1,341), we tested if the difference in educational mobility between siblings was associated with difference in DunedinPACE. This design blocks confounding by all factors shared by siblings in a family.

Results were similar to our primary analysis. The sibling with higher educational mobility tended to have slower DunedinPACE as compared with their less-educated sibling (Offspring cohort Cohen’s d=-0.22, 95%CI=[-0.29,-0.14], p<0.009; Gen3 cohort Cohen’s d=-0.25, 95%CI=[-0.31,-0.18], p<0.001; **Supplemental Table S2**). Again, associations were weaker and not statistically different from zero for the Horvath, Hannum, and PhenoAge clocks. Results for the GrimAge clock, which was developed within the Framingham Heart Study, were similar to those for DunedinPACE. (**Supplemental Table S2**).

### Upwardly-mobile participants and participants with slower pace of aging tended to live longer

We next focused our attention on educational gradients in mortality in the Offspring Cohort (n=1,652; Gen3 participants were not included in this analysis because very few deaths occurred in this younger cohort during the follow-up period). Participants who were more upwardly educationally mobile had lower mortality risk (HR=0.89, 95%CI=[0.81,0.98], p=0.014). In parallel, as previously reported (14), participants with faster DunedinPACE were at higher risk of death than those with a slower DunedinPACE (mortality HR 1.61, 95%CI=[1.49,1.74], p<0.001). All DNA-methylation clocks, with the exception of the Horvath clock, were associated with mortality; effect-sizes were attenuated relative to DunedinPACE with the exception of the GrimAge clock, which was developed to predict mortality in the Framingham sample. Full results are reported in **Supplemental Table S3**.

### A faster pace of aging partly mediated educational gradients in mortality risk

Finally, we tested if differences in DunedinPACE mediated educational gradients in mortality risk. We found that DunedinPACE mediated 50% of the association between educational mobility and mortality risk (indirect effect HR=0.93, 95%CI=[0.90,0.95]). Results were robust to methods that allow relaxation of assumptions about exposure-mediator and mediator-outcome confounding and exposure-mediator interactions (46). Full results are reported in **Supplemental Table S4**.

### Sensitivity analysis

DunedinPACE was measured from blood DNA-methylation data. Blood DNA-methylation patterns are affected by smoking history and the white blood cell composition of the DNA sample (47,48). In turn, these factors may also be related to mortality risk. Therefore, we repeated our analysis including covariate adjustment for these factors. Smoking history was recorded from participant reports; white blood cell composition in the DNA sample was estimated using the algorithms proposed by Houseman and colleagues (49). Covariate adjustment for estimated cell counts and participant reports of smoking history resulted in modest attenuation of some effect-sizes; however, all analyses still showed substantial mediation of educational gradients in mortality risk by DunedinPACE (**Supplemental Table S4**). Full results are reported in **Supplemental Tables S5-S8.**

## DISCUSSION

People with higher levels of education tend to live longer, healthier lives as compared to those with less education (50–52). We analyzed data from three generations of the Framingham Heart Study to test if this educational gradient in healthspan and lifespan could reflect effects of education on the pace of biological aging. Participants who were upwardly mobile in educational terms had slower pace of aging, as measured by the DunedinPACE epigenetic clock, and were less likely to die over the follow-up period. Differences in DunedinPACE accounted for roughly half of the association between educational mobility and mortality risk.

DunedinPACE was developed as a surrogate endpoint for interventions targeting healthy lifespan (53–55). Prior studies have reported associations between education and DunedinPACE, and between DunedinPACE and aging-related disease and mortality (14,15,28,30). Our study is the first to follow individuals across the educational-origins ➔ educational-attainment ➔ pace-of-aging ➔ mortality pathway. The magnitude of education-DunedinPACE associations we report in this sample (r=0.19-0.24) are consistent with population-representative studies in the US, UK, and New Zealand (r=0.17-0.38; (15)) and correspond to a 2-3% slower pace of aging per unit of upward educational mobility (approximately 2 years). In turn, our mediation analysis found that this magnitude of slowing in DunedinPACE corresponded to a roughly 7% reduction in the hazard of mortality, or half of the overall effect of educational mobility. Collectively, these findings contribute evidence that DunedinPACE is a candidate surrogate endpoint for the effects of educational interventions on aging.

A further contribution of our study is evidence that healthy-aging returns to education persist into more recent birth cohorts, amongst whom higher levels of education are more common. Educational gradients in mortality have grown steeper in recent years (56). However, these trends reflect outcomes primarily for the cohorts born across the early-to-middle 20^th^ century. Across these cohorts, the proportion of individuals completing high school and college education increased dramatically (57). Whether the trend of widening educational inequality in healthy aging will persist for later-20^th^ century birth cohorts, for whom rates of high school and college graduation have been more stable, is unknown. We found that effect-sizes for associations between upward educational mobility and slower DunedinPACE were similar for the Offspring and Gen3 cohorts, suggesting that even in the context of relatively high educational attainment, upward mobility continues to yield returns for healthy aging.

We acknowledge limitations. There is no gold standard measure of biological aging (24). We focused on the pace of aging measure DunedinPACE based on three lines of evidence. First, DunedinPACE is predictive of diverse aging related outcomes, including disease, disability, and mortality (14,28,30,32,35). Second, DunedinPACE is associated with social determinants of healthy aging in young, midlife, and older adults (14,15,31,33,34,58). Third, DunedinPACE shows evidence of being modified by calorie restriction (59), an intervention that modifies the basic biology of aging in animal experiments (60). Confidence in results is further supported by the consistency of our findings with those for alternative measurements of biological aging in independent cohorts (61). Finally, our results are robust to known confounds of DNA methylation-based measurements of aging, specifically cell composition of blood samples used to derive DNA and smoking history (48,62).

Isolating effects of education on biological aging is challenging because the level of education people complete is strongly influenced by familial factors and other features of early life environment that are associated with the pace of biological aging (31,58,63–65). We addressed this potential confound in two ways. First, we focused on educational mobility between generations of a family. Second, we conducted analyses comparing siblings within a family. Across these specifications, we found consistent evidence of slower pace of aging in people who were upwardly educationally mobile and who completed more schooling as compared to their siblings. Ultimately, evidence from randomized trials (e.g., (66)) will be needed to confirm if promoting educational attainment slows the pace of aging. The process of social mobility also involves post-education attainments. Studies of mobility, biological aging, and lifespan that consider mobility in terms of income, occupation, and wealth accumulation are needed (67,68).

The Framingham sample is predominantly White-identifying and includes relatively few participants with the lowest levels of educational attainment. In nationally representative datasets from the United States, the difference in mortality risk associated with having attended some college as compared to not is HR=0.66-0.76 (69). In the Framingham data we analyzed, the corresponding effect-size is somewhat smaller (HR=0.81). The lowest levels of educational attainment are not well represented in the Framingham Heart Study, possibly attenuating education-mortality associations. The ultimate result should be to make our estimates of mediation by DunedinPACE conservative. Nevertheless, replication of our results in more diverse cohorts is a priority. Measurement of DNA-methylation in the Offspring Cohort at the 8^th^ wave of measurement, after roughly four decades of follow-up, could induce survival bias. However, we observe similar effect-sizes for associations of educational mobility with DunedinPACE in the Offspring Cohort and the younger Gen 3 cohort, for whom DNA methylation data were generated from samples collected at their second wave of measurement. Moreover, the distribution of educational attainment in Offspring Cohort participants included in our analysis is similar to the distribution at study baseline (wave 1 mean=12.1, SD=2.4; wave 8 mean=12.3, SD=2.5). Thus, selective mortality did not bias the distribution of education in our analysis sample.

The healthier aging of individuals with more education and other social advantages is well established. Our study contributes evidence that an accelerated pace of biological aging is among the mechanisms that account for this inequality. In addition, it suggests that new methods to quantify the pace of aging can provide near-term measures of health impacts for programs and policies designed to promote educational attainment and other socioeconomic assets. Because the pace of aging is variable from young adulthood, measurements such as DunedinPACE can illuminate intervention effects years or decades before aging-related functional deficits and chronic diseases become apparent. Such information can, in turn, help refine efforts to heal health disparities and building aging health equity.

## Supporting information

Graf_FraminghamSocMob_Supplement

Graf_FraminghamSocMob_TableS4

## Data Availability

All data produced in the present study are available upon reasonable request to the authors

## Acknowledgements

This project was supported by US National Institutes of Health Grants R01AG073402, R01AG073207, and R21AG078627. GG is supported by T32ES023772. DWB is a fellow of the CIFAR CBD Network.

## Notes

### Competing Interest Statement

AC, TEM, KS, and DWB are listed as inventors on the Duke University and University of Otago Invention DunedinPACE, which is licensed to TruDiagnostic.

### Author Declarations

The study used only openly available human data from the Framingham Offspring Study obtained from dbGaP (phs000007.v33.p14 and phs000724.v10.p14).

