## Supplementary material for "Educational Mobility, the Pace of Biological Aging, and Lifespan in the Framingham Heart Study": Graf_FraminghamSocMob_Supplement

### Supplemental Methods

**Construction of educational mobility variables**

Educational attainment was measured differently in the Original, Offspring, and Gen3 Framingham cohorts. We harmonized measures of educational attainment and calculated educational mobility following the method used by Liu and colleagues (18).

We converted the educational levels of participants and their parents to Z-scores based on the means and standard deviations of other Framingham Heart Study participants born within five years of the index participant’s year of birth. Mean and standard deviation of educational attainment (in years) are shown in the table below:

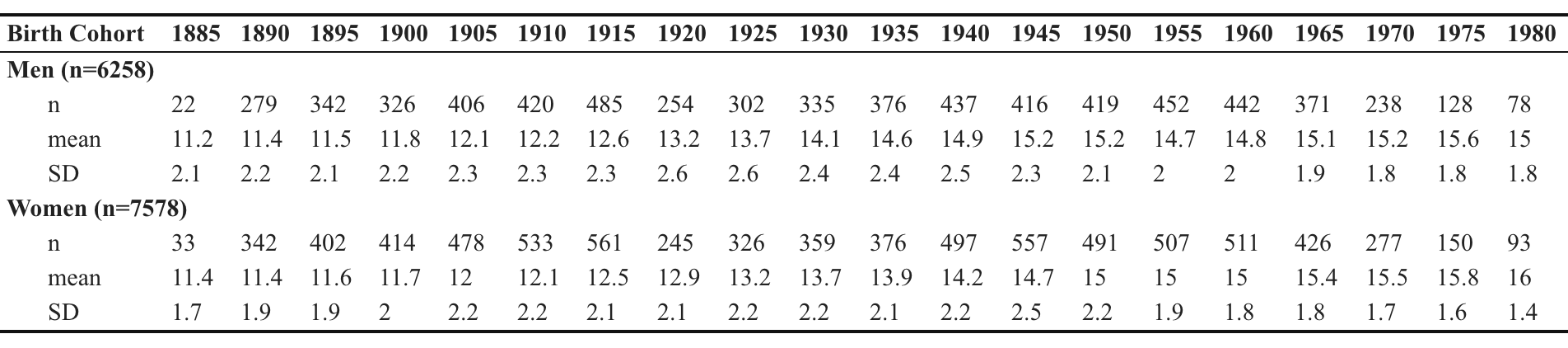

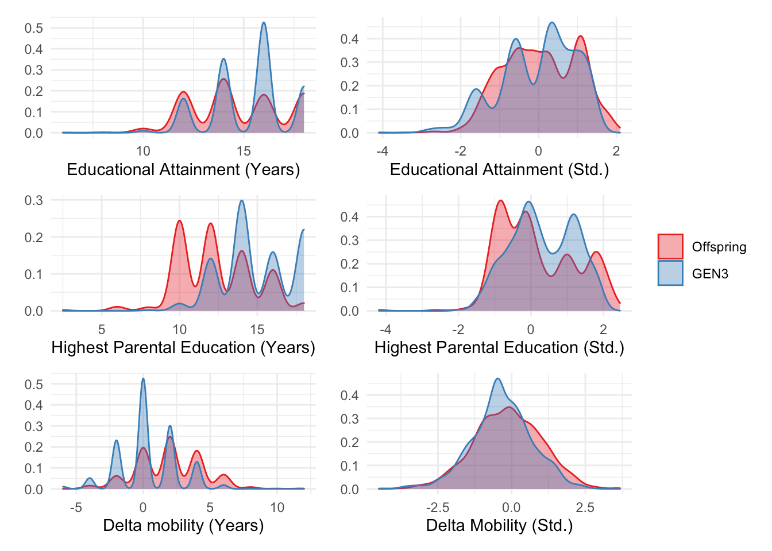
The first two rows of the figure (right) show the distribution of unstandardized and sex- and birth-cohort-standardized educational attainment and parental educational attainment in the Offspring and Gen3 cohorts of the Framingham Heart Study. The last row of the figure shows mobility distribution based on subtracting parents’ raw years of education from that of participants (left), as well as subtracting the standardized measures (right). Both Offspring and Gen3 cohort members were upwardly educationally mobile: Offspring cohort members achieved an average of 3 more years of education than their highest-achieving parent, while Gen3 cohort members achieved an average of 0.4 more years of education than their highest-achieving parent.

In the Offspring cohort for whom comprehensive mortality follow-up was available, participants with a faster Pace of Aging were more likely to die (HR 1.61, 95%CI=[1.49,1.74], p<0.001) than those with a slower Pace of Aging. Effect-sizes were similar to those previously reported (Belsky et al. mortality HR=1.65 [1.51-1.79]).

**Construction of DNA-methylation data**

DNAm was measured from whole-blood buffycoat samples collected at Visit 8 (2005-2008) for the Offspring Cohort and visit 2 (2009-2011) for the Gen3 cohort using the Infinium HumanMethylation450 BeadChip (Illumina). Processing and normalization of DNAm data has been described previously (70). Briefly, data were normalized using the “dasen” method in the ‘wateRmelon’ R package30 and subjected to downstream QC. Samples with missing rate >1% at p<0.01, poor SNP matching to the 65 SNP control probe locations, and outliers by multidimensional scaling techniques were excluded. Probes with missing rate of >20% at p<0.01 were also excluded.

### Supplemental Tables

**Supplemental Table S1. Associations of educational attainment, parental education, and educational mobility with biological aging in Offspring and Gen3 Framingham Study participants.** Effect-sizes are age- and sex-adjusted Pearson's r correlations estimated from linear regression, and are interpretable as the standard-deviation unit increase in biological aging or pace or aging associated with a 1-SD increase in educational attainment or educational mobility. Educational attainment and mobility appear to have modest but statistically significant effects on the Pace of Aging in both Offspring and Gen3 cohorts.

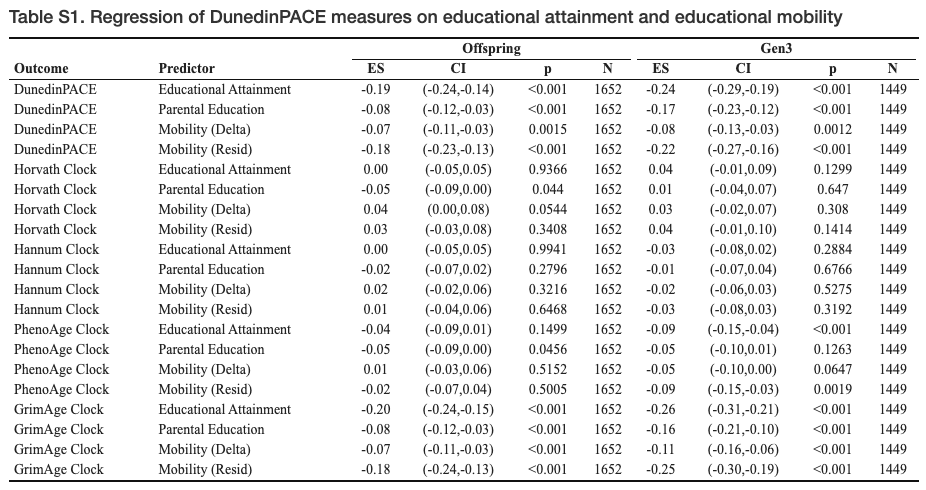

**Supplemental Table S2. Fixed-effects regression of biological aging on education within sibling clusters**. Effect-sizes are interpretable as the standard-deviation unit increase in biological-age advancement or pace of aging associated with a 1-SD increase in educational attainment for among siblings who share the same parents. Because siblings who share the same parents share the same expected educational attainment, results provide further evidence that educational mobility is associated with slower biological aging

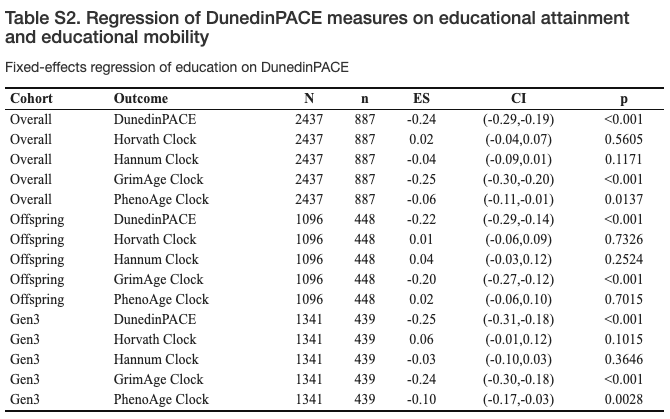

**Supplemental Table S3. Associations of biological aging, educational attainment, and educational mobility with overall survival in the Framingham Offspring cohort.** The table shows age-adjusted hazard ratios estimated from Cox proportional hazards regression. Hazard ratios are interpretable as the change in mortality risk associated with a 1-SD increase in the pace of aging, in educational attainment, or educational mobility. Slower biological aging, educational attainment, and upward educational mobility are associated with overall survival.

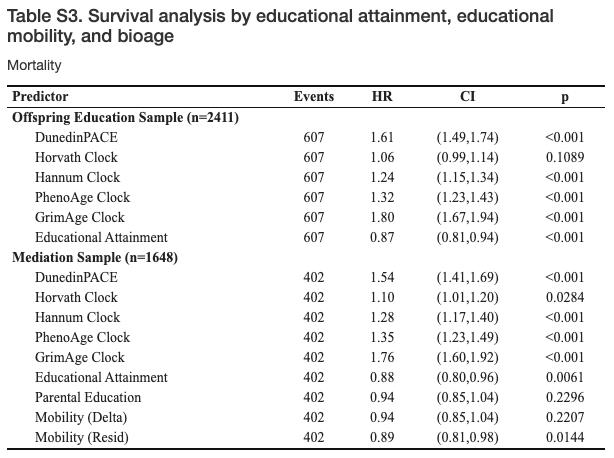

**Supplemental Table S4. Tests of DNA-methylation biological aging measures as mediators of educational gradients in mortality risk.** The table shows results of mediational analysis following the approach of Valeri and Vanderweele (2013). The first table row shows the controlled direct effect (CDE), second row shows the pure natural direct effect (PNDE), third row shows the total natural direct effect (TNDE), fourth row shows the pure natural indirect effect (PNDE), fifth row shows the total natural indirect effect (TNIE), sixth row shows the total effect (TE), last row shows the proportion mediated (PM). For estimates of the controlled direct effect, the value of the mediator (biological-age) is set to zero.Because there is no established gold standard of biological aging, we repeated our primary analysis of the pace of aging measure using four additional DNA-methylation aging “clocks” which have similarly been shown to predict morbidity and mortality in diverse samples, and which have been shown to be sensitive to a range of socioenvironmental exposures. Note that the total effect is equal to the sum of the pure natural direct effect and the total natural indirect effect (PNDE + TNIE), and to the sum of the total natural direct effect and the pure natural indirect effect (TNDE + PNIE).

*See Excel file.*

**Supplemental Table S5. Cell-count-adjusted associations of educational attainment, parental education, and educational mobility with biological aging in Offspring and Gen3 Framingham Study participants.** Effect-sizes are age- and sex-adjusted Pearson's r correlations estimated from linear regression, and are interpretable as the standard-deviation unit increase in pace of aging associated with a 1-SD increase in educational attainment or educational mobility. Educational attainment and mobility appear to have modest but statistically significant effects on the Pace of Aging in both Offspring and Gen3 cohorts.

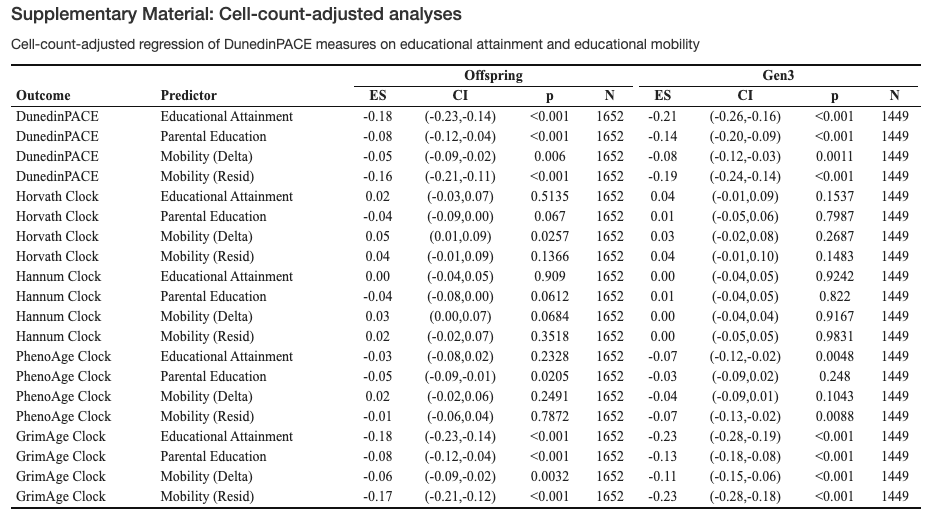

**Supplemental Table S6. Cell-count adjusted associations of biological aging, educational attainment, and educational mobility with overall survival in the Framingham Offspring cohort.** The table shows age-adjusted hazard ratios estimated from Cox proportional hazards regression. Hazard ratios are interpretable as the change in mortality risk associated with a 1-SD increase in the pace of aging, in educational attainment, or educational mobility. Slower biological aging, educational attainment, and upward educational mobility are associated with longer overall survival.

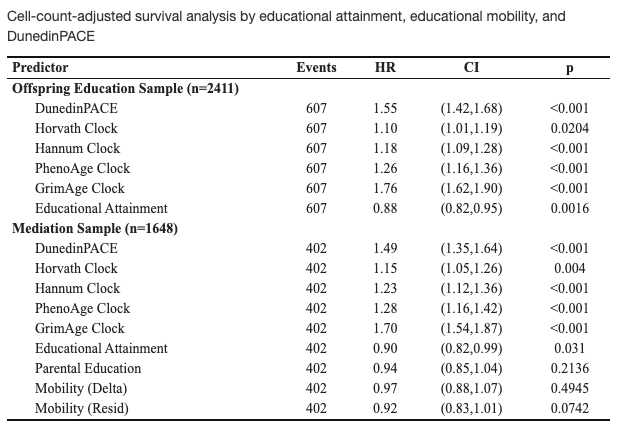

**Supplemental Table S7. Smoking-adjusted associations of educational attainment, parental education, and educational mobility with biological aging in Offspring and Gen3 Framingham Study participants.** Effect-sizes are age- and sex-adjusted Pearson's r correlations estimated from linear regression, and are interpretable as the standard-deviation unit increase in pace of aging associated with a 1-SD increase in educational attainment or educational mobility. Educational attainment and mobility appear to have modest but statistically significant effects on the Pace of Aging in both Offspring and Gen3 cohorts.

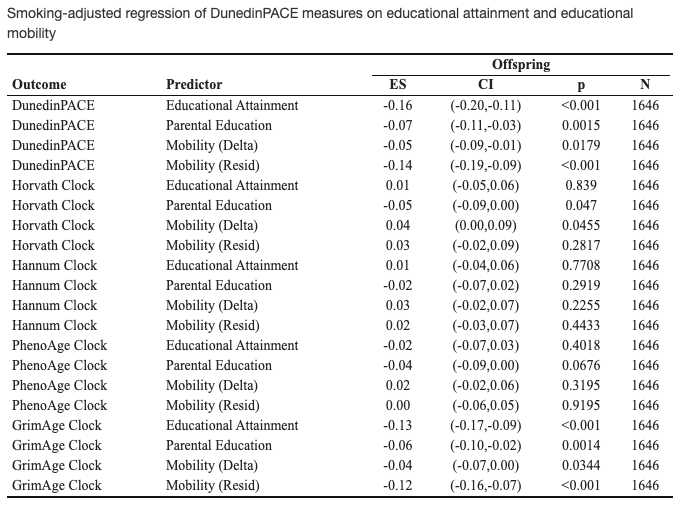

**Supplemental Table S8. Smoking-adjusted associations of biological aging, educational attainment, and educational mobility with overall survival in the Framingham Offspring cohort.** The table shows age-adjusted hazard ratios estimated from Cox proportional hazards regression. Hazard ratios are interpretable as the change in mortality risk associated with a 1-SD increase in the pace of aging, in educational attainment, or educational mobility. Slower biological aging, educational attainment, and upward educational mobility are associated with longer overall survival.

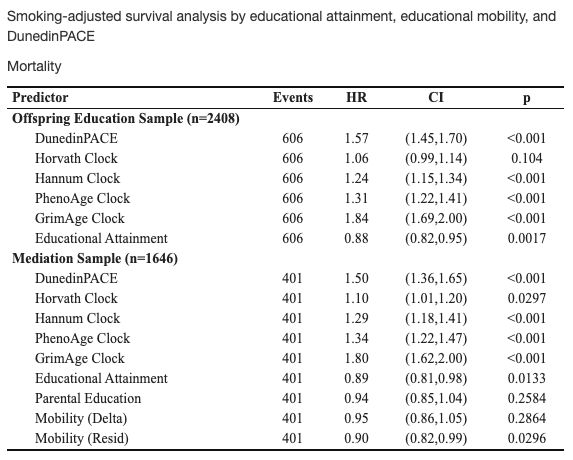

### Supplemental Figures

**Supplemental Figure S1. Intrafamilial education correlation matrices.** Histograms on the diagonal line show the distribution of participants’ own educational attainment and that of their mother, father, and siblings. Pearson’s r correlations are shown above the diagonal line, with corresponding scatterplots and lines of best fit below. Participants’ own educational attainment was moderately correlated with their parents’ educational attainment (r 0.31-0.36) and highly correlated with their siblings’ educational attainment (r 0.73-0.75).

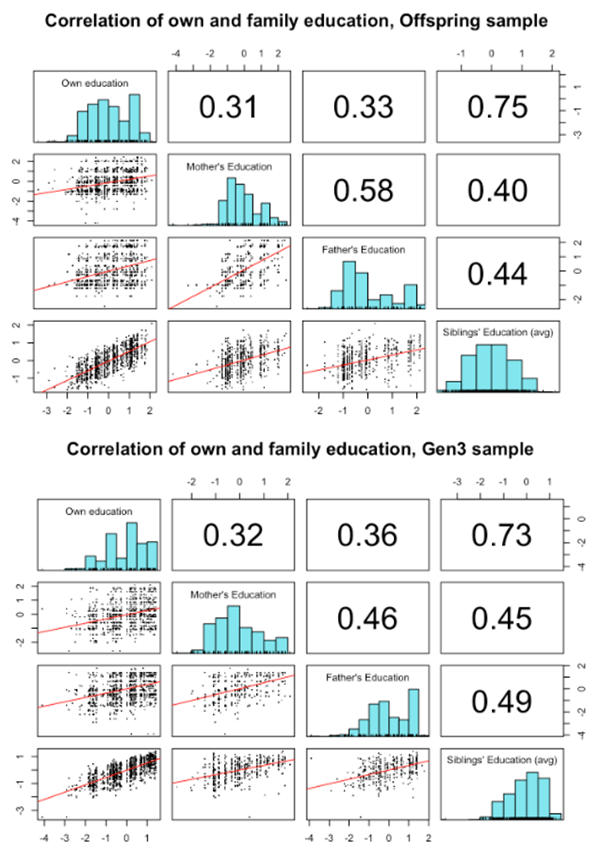

**Supplemental Figure S2. Associations of educational mobility and DunedinPACE by social origins in Offspring and Gen3 Framingham study participants.** This figure shows associations of educational mobility with faster pace of aging, stratified by parental educational attainment. Effect-sizes are Pearson’s r correlations, denominated in standard-deviation units of pace of aging. Observed returns of educational mobility to healthy aging were observed regardless of social origins, with similar effect-sizes across Offspring and Gen3 Framingham study participants.

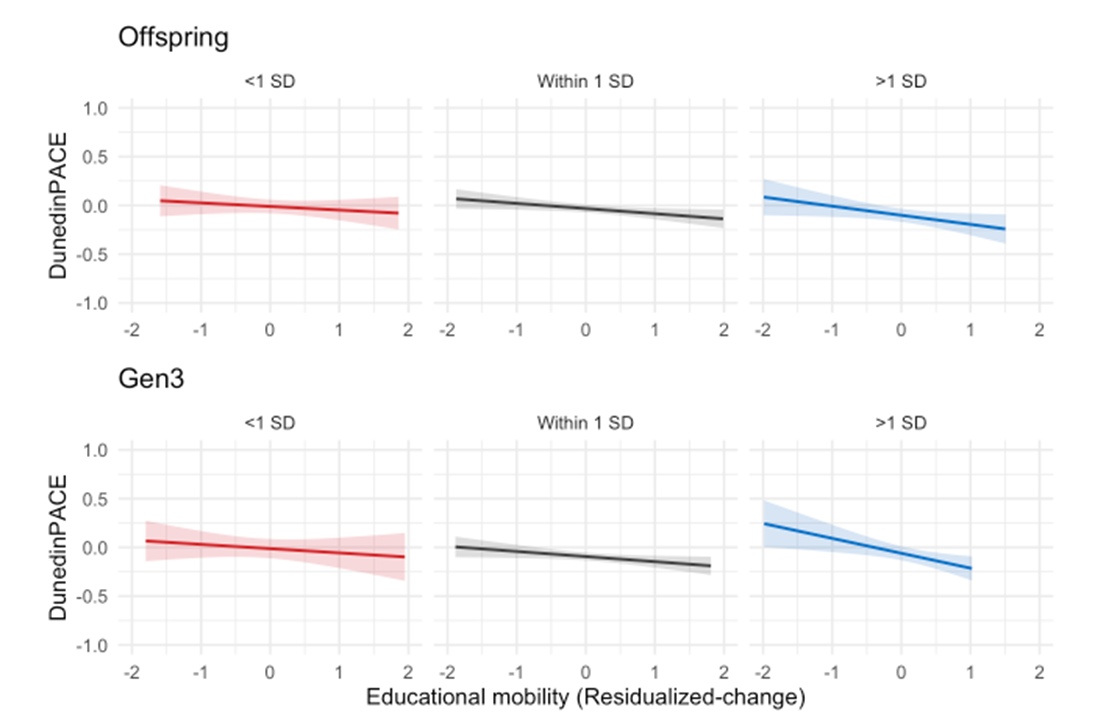
